# Protecting the protectors: Occupational burden and management of work-related musculoskeletal disorders among Cameroonian healthcare professionals and the Sustainable Development Goals

**DOI:** 10.64898/2026.07.30.26359338

**Authors:** Basil Kum Meh, Franklin Chu Buh, Samuel Honoré Mandengue, Orélien Sylvain Mtopi Bopda

## Abstract

Work-related musculoskeletal disorders (WRMSDs) constitute the primary occupational health burden among healthcare professionals (HCPs) globally and directly threaten health workforce sustainability, a critical concern for Sustainable Development Goal 3 (Good Health and Well-being) and SDG 8 (Decent Work and Economic Growth). Despite high prevalence in Cameroon, downstream occupational effects and management strategies across hospital levels remain undocumented. A hospital-based, analytical cross-sectional study was conducted across five referral hospitals in Douala, Cameroon (30 September 2021 to 31 January 2022). A total of 561 HCPs were enrolled by stratified random sampling. The Modified Nordic Musculoskeletal Questionnaire and structured interviews assessed WRMSD occupational effects and management strategies. Data were analysed using IBM SPSS v26; chi-square tests and logistic regression assessed associations (p < 0.05). The 12-month WRMSD prevalence was 83.4% (468/561). WRMSDs caused absenteeism in 83.4% of affected HCPs (χ^2^ = 14.414; p < 0.001), reduced daily activity capacity in 55.6%, and job dissatisfaction in 60.4%; 38.9% were considering a career change. Management was predominantly reactive: fitness training (76.3%), rest (41.4%), and non-steroidal anti-inflammatory drugs (NSAIDs; 37.3%). Only 15.0% received physiotherapy referral, and no hospital had a formal WRMSD protocol. WRMSDs impose a substantial and measurable occupational burden on HCPs in Douala. Current management is reactive, non-standardised, and under-integrated with physiotherapy. Evidence-based, multicomponent prevention programmes, physiotherapy integration, and a national occupational health policy are urgently required to protect the healthcare workforce and advance SDG 3 and SDG 8 targets in Cameroon.

## Introduction

Work-related musculoskeletal disorders (WRMSDs) are conditions of the muscles, tendons, nerves, cartilage, ligaments, and spinal discs that are caused or exacerbated by occupational exposures [1]. Globally, the burden of WRMSDs is substantial: the Global Burden of Disease study identified musculoskeletal disorders as the leading cause of years lived with disability worldwide, affecting an estimated 1.71 billion people [2]. Low back pain alone affected 619 million people in 2020 and is projected to reach 843 million by 2050, with the most dramatic growth expected in Africa [3]. These disorders are the leading driver of absenteeism, lost productivity, and work-related disability across all occupational sectors globally [4,5], representing a direct threat to both SDG 3 (Good Health and Well-being) and SDG 8 (Decent Work and Economic Growth).

In healthcare settings, WRMSDs are disproportionately prevalent. A comprehensive global systematic review reported that the lower back, neck, and shoulder were the most affected body regions across all healthcare professions, with nurses and medical laboratory scientists consistently recording the highest prevalence [6]. A global meta-analysis of nurses reported an overall 12-month WRMSD prevalence of 83.9% [7], while a scoping review of sub-Saharan African (SSA) nurses found lower back pain prevalence ranging from 33% to 90.1% [8].

In SSA, the burden is compounded by severe healthcare workforce challenges. The region carries approximately 25% of the global disease burden while employing only 3% of the world’s health workforce [9]. Projections indicate a critical shortfall of health workers: needs-based modelling estimated a shortage of 6.1 million health workers in the WHO Africa Region by 2030 [10], and as of 2024, Africa had only 46% of the health workers it required, with nearly 943,000 trained professionals unemployed despite critical service understaffing, a systemic paradox of scarcity and waste [11]. Furthermore, the deployment of advanced practice nursing roles has been advocated as a partial solution to primary care shortages in the region [12]. Under these conditions, any factor driving absenteeism, presenteeism, or career attrition constitutes a compounded threat to healthcare delivery capacity and SDG workforce targets.

Beyond their immediate clinical impact, WRMSDs impose major occupational consequences. Kang et al. demonstrated that WRMSDs were independently associated with absenteeism, presenteeism, perceived productivity loss, and work limitations among hospital nurses [7]. Krishnanmoorthy et al., in a systematic review of 19 participatory ergonomic intervention studies, confirmed WRMSDs as the leading driver of sickness absence in nursing populations globally [13]. At the individual level, WRMSD-related presenteeism has been identified as amplifying vulnerability and limiting career advancement, fuelling career dissatisfaction and intent to leave the profession [14], occupational consequences that are particularly alarming in the African context [15].

The management of WRMSDs encompasses multicomponent strategies, including ergonomic education and workplace redesign, physiotherapy-led rehabilitation, pharmacological management, and multidisciplinary occupational health interventions [16,13]. International guidelines recommend non-pharmacological approaches as first-line, with NSAIDs as adjuncts [17]. Multicomponent participatory ergonomic interventions have demonstrated superior effectiveness, achieving significant WRMSD reduction at both six months (OR 1.64; 95% CI 1.12–4.54) and 12 months (OR 2.70; 95% CI 1.52–4.51) post-intervention [13].

In Cameroon, Meh et al. documented an 83.4% WRMSD prevalence in 561 HCPs across five referral hospitals in Douala [18]. A subsequent paper established associations between anthropometric variables and WRMSD occurrence [19]. However, neither study addressed the downstream occupational effects on work attendance and professional functioning, nor the management strategies in use across institutional levels. This gap constitutes the principal novelty of the present work.

This study therefore aimed to: (1) assess the occupational effects of WRMSDs on work quality, attendance, and professional functioning; (2) document and characterise the management strategies employed across five referral hospitals in Douala; and (3) identify variation in management practice across hospital types and professional specialties. Findings are intended to inform occupational health policy and the development of standardised, evidence-based WRMSD protocols in Cameroon and comparable resource-limited settings, in direct support of SDG 3 and SDG 8 targets.

## Materials and methods

### Study design and setting

A hospital-based, quantitative, analytical cross-sectional study was conducted in five publicly designated referral hospitals in Douala, Cameroon (estimated population 4.06 million) [20]. Douala is the economic capital and most populous city of the country. Participant recruitment and data collection ran from 30 September 2021 to 31 January 2022. The five hospitals represented all major levels of the national health pyramid: General Hospital Douala (GHD; Level 4, tertiary), Laquintinie Hospital Douala (LHD; Level 3, secondary), Bonassama District Hospital (BDH), Newbell District Hospital (NBDH), and Nylon District Hospital (NDH) (all Level 1, district). This multi-level design enabled comparative analysis across institutional workload and resource gradients.

### Study population, eligibility, and sample size

The target population comprised all full-time healthcare professionals at the five hospitals, including nurses, medical laboratory scientists (MLS), medical doctors (MDs), physiotherapists (PTs), pharmacists, and allied health professionals. Inclusion criteria were: age 21–60 years; full-time employment for at least six months; and written informed consent. Exclusion criteria were: pre-existing musculoskeletal pathology unrelated to work, current pregnancy, and temporary or locum assignment.

Sample size was calculated using Cochran’s formula (n = z^2^pq/d^2^) with p = 0.85, z = 1.96, and d = 0.05, yielding n ≥ 385. Adjusting for 10% non-response gave a minimum of n = 426. The achieved sample of 561 participants substantially exceeded this threshold. Stratified random sampling was employed, with each hospital constituting a stratum and intra-stratum allocation proportional to staff strength by professional cadre.

### Data collection instruments and procedures

Data were collected through face-to-face structured interviews administered by six trained research assistants. Two validated instruments were used. The Modified Nordic Musculoskeletal Questionnaire (MNQ) [21] assessed 12-month WRMSD prevalence across nine body regions and captured self-reported occupational effects on absenteeism, work ability, job performance, and job satisfaction. A supplementary structured questionnaire documented management strategies received or employed, covering medical consultation, physiotherapy referral, pharmacotherapy, ergonomic interventions, and exercise programmes.

Anthropometric and physiological measurements followed the WHO STEPS protocol [22]: body mass index (BMI) was calculated as weight (kg)/height^2^ (m^2^); blood pressure was measured using a validated automated sphygmomanometer (mean of three readings at five-minute intervals).

### Primary and secondary outcomes

Primary outcomes were: (1) self-reported occupational effects of WRMSDs on absenteeism, reduced working hours, impaired daily activities, job dissatisfaction, intention to change profession, and reduced work ability; and (2) the nature, type, and frequency of management strategies employed. Secondary outcomes were variation in management by hospital type, professional specialty, and age group.

### Statistical analysis

Data were double-entered in Microsoft Excel 2019 and exported to IBM SPSS Statistics v26 (IBM Corp., Armonk, NY, USA). Categorical variables are reported as frequencies and percentages. Chi-square (χ^2^) tests of independence assessed associations between categorical variables; binary logistic regression identified independent predictors of WRMSD-related absenteeism; one-way ANOVA compared continuous variable means across groups. Statistical significance was set at p < 0.05 (two-tailed) with 95% confidence intervals.

### Ethical considerations

Ethical clearance was obtained from the Institutional Review Board (IRB) of the Faculty of Health Sciences, University of Buea (No. 2021/1511-07/UB/SG/IRB/FHS), approved on 28 September 2021 and valid until 28 September 2022. Administrative research authorisation for the three district hospitals was granted by the Littoral Regional Delegation of Public Health (No. 1110/AAR/MINSANTE/DRSPL/BCASS), and separate authorisations were issued by the medical directorates of Douala General Hospital (No. 137/AR/MINSANTE/HGD/DM/09/21) and Laquintinie Hospital Douala (No. 0553/AR/MINSANTE/DHL), and by the directorates of Nylon and New-Bell District Hospitals. Copies of all approval and authorisation documents have been provided to the journal. Recruitment commenced after ethical clearance had been granted, and all data collection fell within the Board’s period of validity. All research involving human participants was conducted according to the principles expressed in the Declaration of Helsinki (2013 revision). Written informed consent was obtained from all participants before enrolment; participation was voluntary and anonymous, and could be withdrawn at any time without consequence.

### Use of generative AI tools

An AI-assisted writing tool (Claude, Anthropic) was used solely to assist with formatting the manuscript to the journal’s requirements and to check for grammatical errors. No AI tool was used to generate, analyse, or interpret scientific data or conclusions, and no AI tool is credited as an author. All authors reviewed the manuscript in full and take full responsibility for the accuracy and integrity of its content.

## Results

### Socio-demographic characteristics

A total of 561 HCPs participated (response rate 100%). The majority were female (52.6%), aged 30–39 years (41.4%), and nurses (62.9%). Most held a bachelor’s degree or equivalent (49.9%). LHD contributed the largest sub-sample (29.8%). A total of 37.8% had fewer than three years of service at their current institution (Table 1).

**Table 1.** Socio-demographic characteristics of study participants (N = 561).

| Variable | Category | n | % |
| --- | --- | --- | --- |
| **Gender** |  |  |  |
|  | Female | 295 | 52.6 |
|  | Male | 266 | 47.4 |
| **Age group** |  |  |  |
|  | 20–29 years | 200 | 35.7 |
|  | 30–39 years | 232 | 41.4 |
|  | 40–49 years | 107 | 19.1 |
|  | ≥50 years | 22 | 3.9 |
| **Specialty** |  |  |  |
|  | Nurse | 353 | 62.9 |
|  | Medical laboratory scientist | 80 | 14.3 |
|  | Medical doctor | 52 | 9.3 |
|  | Physiotherapist | 26 | 4.6 |
|  | Others | 50 | 8.9 |
| **Hospital** |  |  |  |
|  | General Hospital Douala (GHD) | 120 | 21.4 |
|  | Laquintinie Hospital Douala (LHD) | 167 | 29.8 |
|  | Bonassama District Hospital (BDH) | 100 | 17.8 |
|  | Newbell District Hospital (NBDH) | 94 | 16.8 |
|  | Nylon District Hospital (NDH) | 80 | 14.3 |
| **Educational level** |  |  |  |
|  | Secondary | 167 | 29.8 |
|  | Bachelor's degree or equivalent | 280 | 49.9 |
|  | Postgraduate | 114 | 20.3 |
| **Longevity in service** |  |  |  |
|  | <3 years | 212 | 37.8 |
|  | 3–5 years | 136 | 24.2 |
|  | 6–10 years | 98 | 17.5 |
|  | >10 years | 115 | 20.5 |

### Prevalence and body region distribution of WRMSDs

The overall 12-month WRMSD prevalence was 83.4% (468/561). The lower back was the most affected body region (58.8%; n = 330), followed by the neck (52.0%), shoulders (49.6%), upper back (46.7%), knees (42.6%), wrists/hands (38.5%), ankles/feet (35.8%), hips/thighs (31.2%), and elbows (20.3%). By specialty, MLS recorded the highest prevalence (88.8%), followed by nurses (81.9%), physiotherapists (80.8%), and medical doctors (78.8%). The association between specialty and WRMSD prevalence was statistically significant (p = 0.001). Risk factors most strongly associated with WRMSDs included working in a static position (90.6%), working with vibrating objects (89.2%), and job stress (87.3%).

### Occupational effects of WRMSDs on work quality and professional functioning

Table 2 summarises the self-reported occupational effects of WRMSDs. Absenteeism was reported by 83.4% (468/561) of affected HCPs (χ^2^ = 14.414; p < 0.001). Over half (55.6%) reported reduced ability to perform normal daily activities, 52.4% reported a reduction in working hours, and 60.4% expressed job dissatisfaction attributable to WRMSDs. A substantial proportion (38.9%) indicated they were considering a career change. HCPs at NDH reported the highest proportion of significantly reduced work ability (67.0%; p = 0.048). The occupational impact was most pronounced among nurses and MLS.

**Table 2.** Self-reported occupational effects of WRMSDs on work quality and professional functioning (N = 561).

| Occupational effect | Response | n | % |
| --- | --- | --- | --- |
| Absenteeism attributable to WRMSDs<sup>a</sup> | Yes | 468 | 83.4 |
|  | No | 93 | 16.6 |
| Reduced ability to perform daily activities | Yes | 312 | 55.6 |
|  | No | 249 | 44.4 |
| Reduction in working hours | Yes | 294 | 52.4 |
|  | No | 267 | 47.6 |
| Job dissatisfaction attributed to WRMSDs | Yes | 339 | 60.4 |
|  | No | 222 | 39.6 |
| Considering career change due to WRMSDs | Yes | 218 | 38.9 |
|  | No | 343 | 61.1 |
| Prevented from performing normal daily activities | Yes | 302 | 53.8 |
|  | No | 259 | 46.2 |
| Significantly reduced work ability at NDH<sup>b</sup> | Yes | 67 | 67.0 |
|  | No | 33 | 33.0 |

<sup>a</sup> Chi-square test: $\chi^2 = 14.414$ ; $p < 0.001$ . <sup>b</sup> $p = 0.048$ (chi-square). NDH: Nylon District Hospital. WRMSDs: work-related musculoskeletal disorders.

### Management strategies for WRMSDs

Table 3 presents the management strategies employed. Of 468 WRMSD-positive HCPs, 300 (53.5%) consulted a medical doctor. Management was predominantly reactive and non-specific: fitness training and exercise was the most commonly documented general strategy (76.3%), followed by rest or reduced working hours (41.4%) and ergonomic education (23.2%). Only 15.0% received a physiotherapy referral. Among those referred (n = 45), therapeutic massage (22.3%) and supervised exercise therapy (19.3%) were the primary physiotherapy modalities. No hospital had a formal, written institutional WRMSD prevention or management protocol.

**Table 3.**
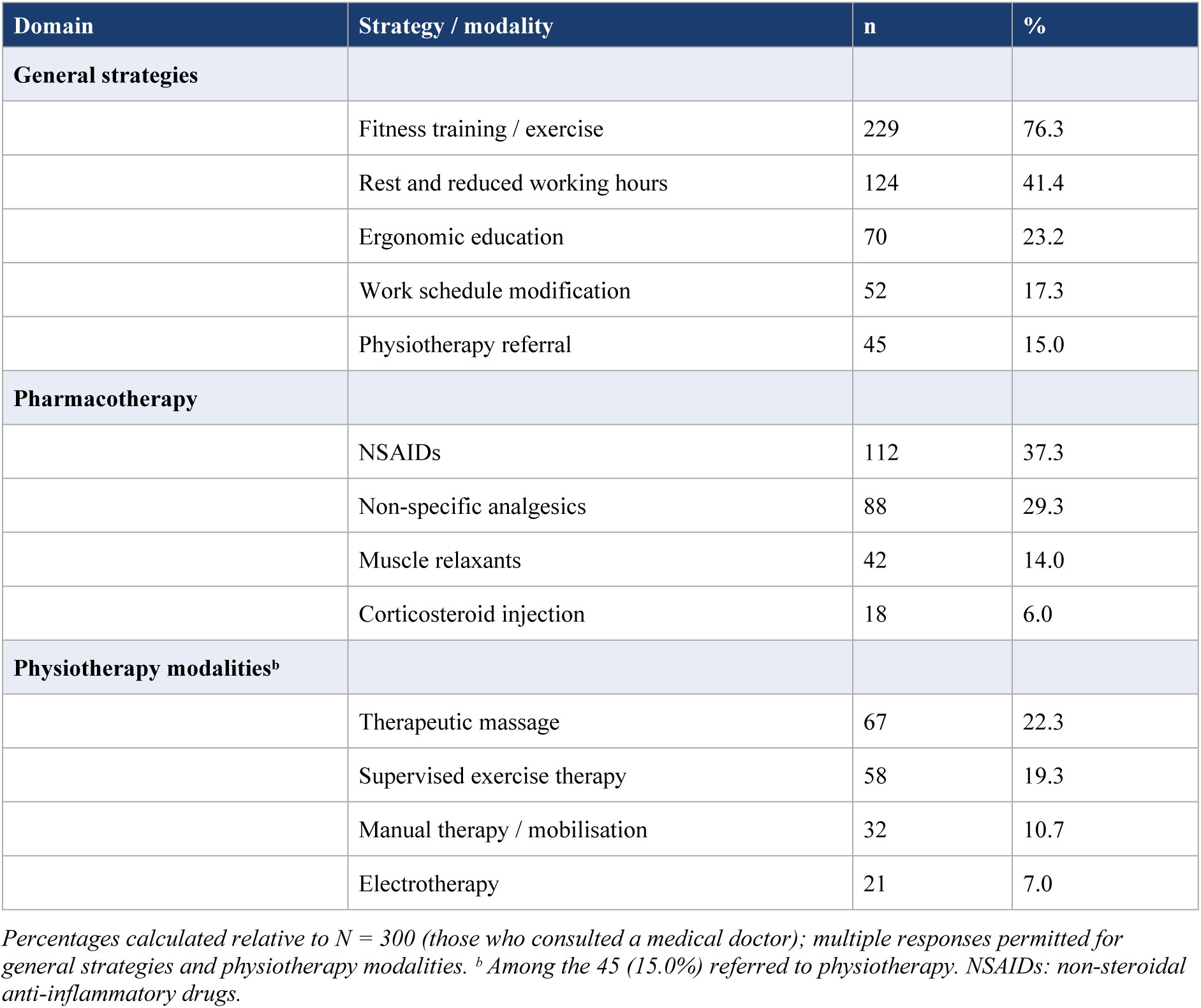
Management strategies employed for WRMSDs among HCPs who consulted a medical doctor (N = 300).

| Domain | Strategy / modality | n | % |
| --- | --- | --- | --- |
| **General strategies** |  |  |  |
|  | Fitness training / exercise | 229 | 76.3 |
|  | Rest and reduced working hours | 124 | 41.4 |
|  | Ergonomic education | 70 | 23.2 |
|  | Work schedule modification | 52 | 17.3 |
|  | Physiotherapy referral | 45 | 15.0 |
| **Pharmacotherapy** |  |  |  |
|  | NSAIDs | 112 | 37.3 |
|  | Non-specific analgesics | 88 | 29.3 |
|  | Muscle relaxants | 42 | 14.0 |
|  | Corticosteroid injection | 18 | 6.0 |
| **Physiotherapy modalities<sup>b</sup>** |  |  |  |
|  | Therapeutic massage | 67 | 22.3 |
|  | Supervised exercise therapy | 58 | 19.3 |
|  | Manual therapy / mobilisation | 32 | 10.7 |
|  | Electrotherapy | 21 | 7.0 |

Percentages calculated relative to N = 300 (those who consulted a medical doctor); multiple responses permitted for general strategies and physiotherapy modalities. <sup>b</sup> Among the 45 (15.0%) referred to physiotherapy. NSAIDs: non-steroidal anti-inflammatory drugs.

NSAIDs were the most commonly prescribed pharmacological agents (37.3%), followed by non-specific analgesics (29.3%), muscle relaxants (14.0%), and corticosteroid injections (6.0%). Management varied significantly by hospital and specialty: GHD workers were significantly more likely to receive ergonomic education than those at district-level hospitals (p = 0.031). Management type differed significantly by age group (χ^2^ = 9.83; p = 0.020), with older HCPs (>40 years) more reliant on fitness and exercise.

## Discussion

This study provides the first comprehensive assessment of the occupational effects and management of WRMSDs across multiple hospital levels and professional specialties in Cameroon. Three overarching findings emerge: (1) WRMSDs affect 83.4% of HCPs and impose statistically significant occupational consequences; (2) absenteeism, reduced work capacity, job dissatisfaction, and career attrition intent directly threaten the stability of an already strained health workforce, undermining SDG 3 and SDG 8 targets; and (3) current management is reactive, pharmacotherapy-dominant, and critically under-integrated with physiotherapy and ergonomic prevention.

### Prevalence and body region distribution: physiological and occupational basis

The 12-month WRMSD prevalence of 83.4% is consistent with the current global evidence base. Jacquier-Bret and Gorce confirmed that the lower back, neck, and shoulder are the most frequently affected body regions across all healthcare professions worldwide, with nurses and laboratory scientists consistently recording the highest burden [6]. A global meta-analysis of nurses found a pooled 12-month prevalence of 83.9%, closely corroborating our findings [7]. In SSA specifically, Kgakge et al. reported lower back pain prevalence ranging from 33% to 90.1% across 25 studies on nurses, a range within which the present lower back finding (58.8%) comfortably falls [8].

The physiological basis for the lower back’s disproportionate burden is well established. The lumbar spine carries the majority of the body’s axial load during standing, bending, and lifting. In healthcare settings, patient-handling tasks impose compressive forces on lumbar intervertebral discs that substantially exceed safe biomechanical thresholds when performed repeatedly without assistive equipment [1]. Sustained static posture activates continuous low-level muscle contractions, impeding local blood flow, reducing oxygen delivery to postural muscles, and accelerating intramuscular metabolite accumulation, driving pain sensitisation and neurogenic inflammation [23,24]. This mechanism directly explains why working in a static position (90.6%) was the most prevalent risk factor in this cohort.

The elevated WRMSD prevalence among MLS (88.8%) is noteworthy. MLS workers maintain prolonged static postures at laboratory benches, perform repetitive fine motor tasks, and work under sustained visual concentration, a combination imposing compound biomechanical loading across the neck, shoulders, and wrists. This replicates data from Fikre et al., who documented high WRMSD prevalence among Ethiopian laboratory professionals attributable to static bench postures [25]. Felix and Siew, in a study of registered nurses in Malaysian hospitals, similarly found inadequate institutional risk management to be the dominant organisational failure contributing to WRMSD persistence, a finding directly applicable to the Douala context [26]. Branca et al., in a narrative review synthesising epidemiological evidence from 2000 to 2025, confirmed that static posture, repetitive motion, forceful exertion, and psychosocial job demands consistently constitute the principal WRMSD risk determinants across healthcare sectors [27].

### Occupational effects: a public health concern in the context of SDG workforce targets

The association between WRMSDs and absenteeism (χ^2^ = 14.414; p < 0.001) directly replicates evidence from Kang et al., who demonstrated independent associations between WRMSDs and all four dimensions of productivity loss in a study of 607 hospital nurses [7]. In SSA, a study at a Ghanaian tertiary hospital confirmed musculoskeletal disorders as the most frequently cited cause of absenteeism [28].

The finding that 60.4% of affected HCPs reported job dissatisfaction and 38.9% were considering a career change is, in the context of the African health workforce crisis, a critical public health finding. The Lancet Commission on Health Workforce estimated that SSA has only 3% of the global health workforce to manage 25% of the global disease burden [9]. WHO AFRO projected a shortage of 5.6 million health workers across the WHO Africa Region by 2030, with 37 countries currently on the WHO Health Workforce Support and Safeguards List [29]. Systemic failures in employment, retention, and distribution rather than training alone have been identified as the defining challenge [11]. Pieterse demonstrated that concurrent skilled health worker shortages and underemployment form a compounding dynamic in SSA [15]. Any WRMSD-driven attrition through absenteeism, reduced hours, or career departure could meaningfully disrupt healthcare delivery capacity and delay achievement of SDG 3.8 (universal health coverage).

The physiological dimension of presenteeism deserves specific emphasis. The finding that 52.4% of HCPs reduced their working hours and 55.6% reported impaired daily activities while remaining employed indicates widespread functional impairment. Physiologically, healthcare professionals working with unmanaged chronic musculoskeletal pain experience sustained activation of the sympathoadrenal system, with elevated cortisol and catecholamines impairing executive function, psychomotor speed, and fine motor control [30], directly affecting the clinical competencies required for safe patient care. Kolovou et al. observed that WRMSD-related presenteeism amplifies nursing staff vulnerability and may limit career advancement opportunities [14].

The concentration of significantly reduced work ability at NDH (67.0%; p = 0.048), a Level 1 district facility, illustrates a critical paradox: those at greatest occupational risk receive the least infrastructure support. District-level facilities in Cameroon typically lack patient-handling equipment, ergonomic workstations, and occupational health services, intensifying physical loading while offering no compensatory preventive support. This finding aligns with Kgakge et al.’s SSA scoping review, which identified structural and staffing deficits at primary-level facilities as independent amplifiers of WRMSD functional impact [8].

### Management: a reactive culture that contradicts international evidence

The management landscape revealed by this study fundamentally contradicts the current international evidence base. The predominance of fitness training (76.3%) and rest (41.4%) as general strategies, without integration into structured physiotherapy-based or ergonomic rehabilitation programmes, reflects a reactive, symptom-driven culture rather than a prevention-oriented one. The use of rest as an isolated strategy for musculoskeletal pain is explicitly discouraged by contemporary guidelines: sustained rest promotes muscle deconditioning, reduces functional capacity, and may prolong pain sensitisation through disuse-related neuroplastic changes in the spinal dorsal horn [31,16].

Multicomponent interventions integrating supervised exercise, ergonomic education, and organisational workplace modifications are substantially more effective than single-component approaches. Krishnanmoorthy et al., in a systematic review of 19 studies, demonstrated that multicomponent participatory ergonomic interventions reduced WRMSDs significantly at both six months (OR 1.64; 95% CI 1.12–4.54; p = 0.011) and 12 months (OR 2.70; 95% CI 1.52–4.51; p < 0.001) [13]. The WHO guideline for chronic primary low back pain explicitly recommends supervised exercise programmes and cognitive behavioural therapy before pharmacological agents [17]. Gkougkoulias et al. confirmed significant reductions in musculoskeletal pain intensity and absenteeism with multicomponent ergonomic programmes [24]. Most recently, Adebiyi et al. reported pain reductions of 38% in the neck and 37% in the hand and wrist through combined ergonomic redesign and supervised physical activity, outcomes achievable only through simultaneous delivery of both components [32].

The very low physiotherapy referral rate (15.0%) is the most clinically alarming finding in this management analysis. Physiotherapy-led rehabilitation encompassing supervised exercise therapy, manual therapy, and functional rehabilitation is the recognised first-line non-pharmacological intervention for WRMSDs [16,33]. Physiologically, supervised exercise restores musculotendinous flexibility, rebuilds stabilising muscle endurance, normalises movement patterns, and stimulates endogenous opioid and serotonin pathways that reduce pain sensitisation [34]. Manual therapy specifically activates supraspinal descending pain inhibition via the periaqueductal grey matter, as elaborated in mechanistic modelling by Bialosky et al. [35]. Pain neuroscience education (PNE) has been shown in a systematic review of 15 randomised controlled trials to significantly improve pain, disability, and psychosocial outcomes in chronic musculoskeletal pain [36]. The near-absence of physiotherapy integration reflects both limited deployment of occupational physiotherapy services and a systemic knowledge gap among hospital managers, a gap explicitly identified by Harithasan et al. [37].

The pharmacotherapy picture, NSAID prescription in 37.3%, analgesics in 29.3%, is not without physiological risk when used in isolation. Chronic NSAID use without concurrent rehabilitation exposes HCPs to gastrointestinal, renal, and cardiovascular adverse effects. A 2025 analysis of over 100,000 NSAID-related adverse event reports confirmed gastrointestinal injury as the most common adverse outcome, and cardiovascular events as the most life-threatening [38]. Anderson et al. and Karran et al. both emphasise that NSAIDs should serve as short-term adjuncts within a multicomponent management plan, not as standalone treatment [39,31].

The significant institutional gradient in ergonomic education, with GHD most likely to provide it (p = 0.031), reveals a structural inequity: district-level facilities, where workers face the highest relative physical workloads and the fewest resources, received the least preventive support. This aligns with broader literature on preventive intervention distribution inequity in healthcare systems [40] and underscores the need for equity-focused occupational health strategies within SDG 10 (Reduced Inequalities) frameworks.

## Conclusion

WRMSDs impose a major, measurable, and growing occupational burden on healthcare professionals across referral hospitals in Douala, Cameroon. They are robustly associated with absenteeism, reduced work capacity, job dissatisfaction, and career change intent, consequences that, in the context of the African health workforce crisis, represent a direct threat to healthcare system sustainability and to the achievement of SDG 3 and SDG 8 targets. Current management is reactive, non-standardised, pharmacotherapy-dominant, and critically under-integrated with physiotherapy. No hospital surveyed had a formal WRMSD prevention or management protocol.

Urgent, evidence-based action is required: multicomponent ergonomic and physiotherapy-integrated prevention programmes; a national occupational health policy specifically addressing WRMSDs in hospital settings; institutional protocol development with management accountability; investment in assistive patient-handling equipment, particularly at district-level facilities; and specialty-specific interventions targeting nurses and medical laboratory scientists, who bear the highest burden. These actions are not only clinically imperative but are essential enablers of SDG 3, SDG 8, and SDG 10 in the Cameroonian context.

## Abbreviations

BMI: body mass index
BDH: Bonassama District Hospital
GHD: General Hospital Douala
HCP: healthcare professional
LHD: Laquintinie Hospital Douala
MLS: medical laboratory scientist
MNQ: Modified Nordic Musculoskeletal Questionnaire
NBDH: Newbell District Hospital
NDH: Nylon District Hospital
NSAIDs: non-steroidal anti-inflammatory drugs
OR: odds ratio
PNE: pain neuroscience education
PT: physiotherapist
SDGs: Sustainable Development Goals
SSA: sub-Saharan Africa
WHO: World Health Organization
WRMSDs: work-related musculoskeletal disorders

## Author contributions

Basil Kum Meh: Conceptualization, Methodology, Formal analysis, Investigation, Data curation, Writing – original draft, Visualization. Franklin Chu Buh: Investigation, Writing – review & editing. Samuel Honoré Mandengue: Supervision, Writing – review & editing. Orélien Sylvain Mtopi Bopda: Supervision, Writing – review & editing, Project administration.

## Data availability

All relevant data are within the manuscript. The minimal underlying dataset cannot be made publicly available without restriction because it contains information collected from identifiable healthcare institutions, and the conditions of ethical approval limit onward distribution. Requests for access to the de-identified minimal dataset should be addressed to the Institutional Review Board, Faculty of Health Sciences, University of Buea, quoting application number 1511-07. The Board is a non-author body and will consider requests from any qualified researcher on the same terms.

## Acknowledgments

The authors sincerely thank all 561 healthcare professionals across the five hospitals for their time and cooperation. Statistical support from Mr Ferdinand Tingom and the Ferdsilinks group is gratefully acknowledged. Administrative support from the Littoral Regional Delegation of Public Health and all five hospital administrators is appreciated.

